# Evaluating the potential of respiratory-sinus-arrhythmia biofeedback for reducing physiological stress in adolescents with autism: a protocol for a randomized controlled study

**DOI:** 10.1101/2021.02.27.21252570

**Authors:** Anoushka Thoen, Jean Steyaert, Kaat Alaerts, Tine Van Damme

## Abstract

**Background:** Prior evidence points towards lower cardiac vagal modulation in individuals with Autism Spectrum Disorder (ASD) as compared to control groups. A cross-sectional phase in this study will gather more evidence concerning this topic. A longitudinal phase will explore the efficacy of a biofeedback intervention based on Respiratory Sinus Arrhythmia (RSA) in adolescents with ASD. Finally, a feasibility study will focus on a non-supervised RSA biofeedback intervention in this population.

**Methods:** The cross-sectional phase includes the comparison of adolescents with ASD (n=38) and age and gender matched typically developing peers. A standardized assessment will be used which contains physiological, cortisol and behavioral measurements. The longitudinal phase contains a randomized, single-blinded and sham-controlled design to determine the efficacy of supervised RSA biofeedback in adolescents with ASD (n=128). A follow-up phase of 5 weeks is included to evaluate the presence of retention effects. During the latter, a feasibility study will focus on a non-supervised intervention (n=62). Assessments as described previously are scheduled after the intervention and the follow-up phase.

**Discussion:** First, more conclusive evidence will be provided for the presence of lower cardiac vagal modulation in adolescents with ASD as well as the association between these lower values and physiological and behavioral indices. Second, the supervised intervention in adolescents with ASD is hypothesized to upregulate this cardiac vagal modulation and positively change behavioral and physiological parameters. Third, evidence regarding the feasibility and acceptability of a non-supervised intervention may open novel avenues for home-based interventions in this population.

**Study registration:** ClinicalTrials.gov, NCT04628715. Registered on 13 November 2020.

**Declarations:** *Funding:* Funding is provided by the Marguerite-Marie Delacroix foundation with grant number GV/B-363. The funder will not have any role in any part of this study.

*Competing interests:* The authors declare that they have no conflict of interest.

*Availability of data and material:* All data from the participants will be de-identified and provided with a unique code. The coded data will be stored for 20 years in secured databases of the Research Group for Adapted Physical Activity and Psychomotor Rehabilitation, protected by the KU Leuven and will only be accessible by researchers of collaborating labs. The key to the coded data will be stored securely and confidentially in a separate electronic file. The saliva samples will be stored under appropriate conditions during the study at the biobank of the KU Leuven and will be destroyed afterwards. Participants waive any intellectual property rights on findings that might result from the analysis of their saliva samples.

*Code availability:* Not applicable.

*Author’s contributions:* All authors contributed to the writing of this manuscript and the grant proposal. Anoushka Thoen leads the study and data management under supervision of Tine Van Damme, who provided facilities and equipment. All authors read and approved the final manuscript.

*Ethics approval:* Ethical approval for this study was granted by both the Ethics Committee UPC KU Leuven on July 2^nd^ 2020 (ref. EC2020-541, version 2.0) and the *Ethics Committee Research UZ/KU Leuven on October 20*^*th*^ *2020 (ref: S*64219, version 1.0).

*Consent to participate:* Parents of the participants should provide informed consent in addition to the informed assent provided by the participants themselves. Participants may withdraw consent and participation at any time. The participant’s request to withdraw from the study will always be respected and reasons to withdraw are not obliged to be mentioned. The sponsor of this study is KU Leuven (Oude Markt 13, 3000 Leuven, Belgium) and will have no role in any part of this study.

*Consent for publication:* Authorship to publications will be determined in accordance with the requirements published by the International Committee of Medical Journal Editors and in accordance with the requirements of the respective peer-reviewed medical journal.

*Amendments:* Every substantial adjustment to the protocol will be communicated to the Ethics Committee UPC KU Leuven and the *Ethics Committee Research UZ/KU Leuven as an amendment to the protocol. Only after approval of this amendment, the adjustments can be implemented and communicated to the researchers and participants*.

*Auditing:* The researchers will permit study-related monitoring, audits, Ethical Committee review and regulatory inspection, providing direct access to all related source data/documents.

*Dissemination of study results:* The results of this study will be used for publication in peer-reviewed journals. There will also be a general dissemination of the study results for the participants and personal results will be provided upon request.

## Background and rationale

In the last decade, research concerning stress-mechanisms in individuals with autism spectrum disorder (ASD) has increased exponentially. This neurodevelopmental disorder is characterized by problems in social interaction and communication and the presence of repetitive, stereotyped behaviors, activities and interests (American Psychiatric Association, 2013). In addition, children and adolescents with ASD report higher rates of stress as compared to typically developing (TD) peers (Bishop-Fitzpatrick et al., 2017; Groden, 2006). Especially in the adolescent period, which is characterized by rapid changes across several developmental domains (e.g. social context, affect regulation and behavior), individuals with ASD are hypothesized to be more vulnerable for stress-related difficulties (Corbett & Simon, 2014).

Researchers recently explored the contributing role of the autonomic nervous system in psychiatric disorders, including ASD (Benevides & Lane, 2015; Condy et al., 2019; Ellenbroek & Sengul, 2017; Lydon et al., 2016; Patriquin et al., 2019). One of the most commonly adopted physiological parameters for assessing autonomic nervous system functioning in psycho-physiological research is based on the assessment of heart rate variability (HRV). The heart’s pacemaker is controlled by both the sympathetic and the parasympathetic nervous system, which results in the natural variation of the interval between heartbeats. One important component of HRV is respiratory sinus arrhythmia (RSA). This component is solely mediated by the parasympathetic subsystem through direct projections of the vagus nerve from the brain stem to the heart and is therefore also known as an index of cardiac vagal modulation. The term RSA is adopted since it defines the phenomenon of the relation between heart rate variability and the respiratory cycle (Shaffer et al., 2014). This parameter is typically identified in the high-frequency component of HRV (HF-HRV), which ranges between 0.15 and 0.4 Hz, and by the root mean square of successive differences (RMSSD) of the interbeat intervals (Shaffer & Ginsberg, 2017).

The clinical importance of RSA has been reported in a variety of studies demonstrating higher RSA to be associated with higher cognitive abilities, effective social behavior, fewer internalizing symptoms (e.g. anxiety and depression) and appropriate emotion regulation (Beauchaine, 2015; Beauchaine et al., 2019; Fiskum, 2019). Moreover, lower RSA has been linked to conduct problems, trait hostility, anxiety disorders and depression (Beauchaine et al., 2019; Graziano & Derefinko, 2013; Patriquin et al., 2015). Another important aspect of RSA is its positive association with appropriate social behavior as described by Porges’ polyvagal theory (Porges, 2011). The latter theory describes a parasympathetically driven social engagement system, which promotes the activation of interconnected craniofacial nerves and their associated motor behaviors. This way, a calm state – as induced by a heightened parasympathetic drive – is associated with prosocial behaviors including the making of eye contact, appropriate vocalizations and socially oriented head turning (Muscatello et al., 2021). Recent research indicated that TD children and adolescents demonstrated more prosocial behavior when higher RSA values were present (Corbett et al., 2019; Muscatello et al., 2021).

With respect to children and adolescents with ASD, the majority of studies have demonstrated lower levels of RSA in individuals with ASD during various activities (Benevides & Lane, 2015; Condy et al., 2019; Ellenbroek & Sengul, 2017; Patriquin et al., 2019). This was confirmed in a recent meta-analysis, where statistically significant lower RSA-values were found in individuals with ASD during baseline measurements, social stress tasks, social debriefing tasks and cognitive tasks (Cheng et al., 2020). Lower RSA levels in children and adolescents with ASD have been associated with higher levels of autism-specific symptoms such as social problems, internalizing problems, anxiety, emotional control issues, more severe visual and/or auditory sensory problems and repetitive and restrictive behaviors (Benevides & Lane, 2015; Condy et al., 2019; Ellenbroek & Sengul, 2017; Patriquin et al., 2019). However, given methodological differences, multiple researchers have recommended further research on ANS functioning in individuals with ASD and how it deviates from TD peers (Cheng et al., 2020; Corbett et al., 2019; Lory et al., 2020; Lydon et al., 2016; Muscatello et al., 2021). Therefore, a cross-sectional phase is included in this study in which a standardized stress-provoking assessment will be used in adolescents with and without ASD.

Considering the prior observations of lower RSA levels in individuals with ASD, it would be of high clinical relevance to develop and validate an intervention that specifically targets the upregulation of RSA-values – a measure of cardiac vagal modulation – in individuals with ASD (Patriquin et al., 2019). Prior HRV-RSA biofeedback studies have shown that an increase of HRV was causally related to several positive effects on psychological parameters. These studies were performed in distinct clinical populations of all ages and variable medical and psychological conditions (Chudleigh et al., 2019; Darling et al., 2019; Gevirtz, 2013; Kennedy & Parker, 2019; Knox et al., 2011; Lehrer et al., 2020; Lehrer & Gevirtz, 2014; Thabrew et al., 2018). In this protocol, individuals practice to breath at their resonant frequency, which can be found between 4.5 and 7 breaths per minute. Breathing at this frequency will maximize HRV oscillations due to the entrainment of two physiological mechanisms affecting HRV, namely the influence of the baroreflex and of the breathing pattern (Fiskum, 2019). For more information concerning the hypothesized working mechanisms, see Lehrer and Gevirtz (2014). Only one recent study provided evidence of feasibility of a similar intervention in a small sample (n = 7) of children and adolescents (10-15 years) with ASD (Goodman et al., 2018). The current study will encompass a large-scale randomized-controlled trial, in which a representative sample of adolescents with ASD will undergo a supervised RSA based biofeedback training to test its efficacy for enhancing RSA-levels and positively changing associated behavioral symptoms and physiological parameters as determined at the cross-sectional phase. Finally, a feasibility study regarding a non-supervised RSA based biofeedback intervention will be implemented for half of the adolescents with ASD after the supervised intervention part (see Figure 1).

**Fig. 1.**
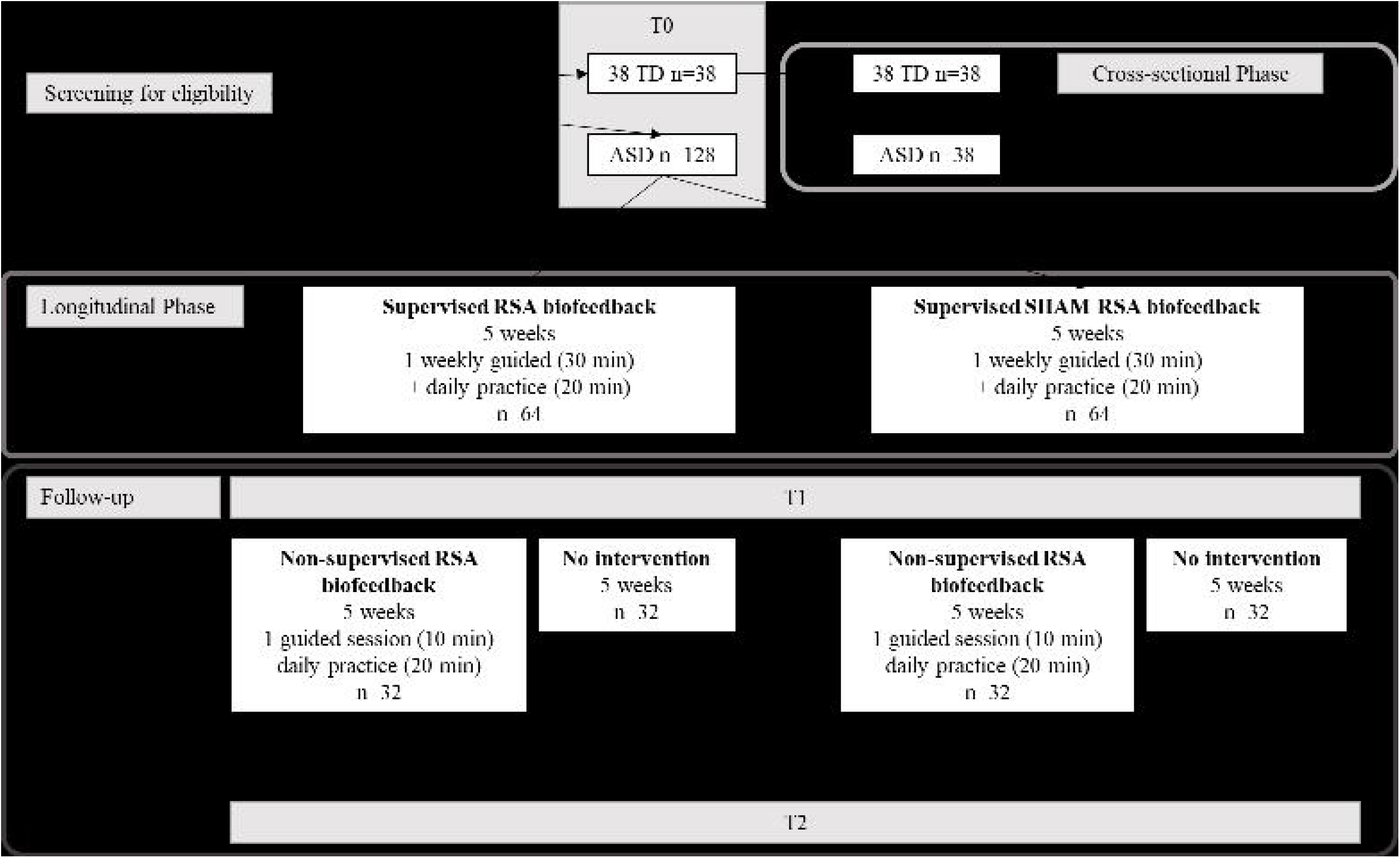
Study Flow Diagram

### Specific aims and hypotheses

This study consists of a cross-sectional and a longitudinal phase, each containing one research objective. An overview of the complete study design is presented in Figure 1 and Table 1.

**Table 1.**
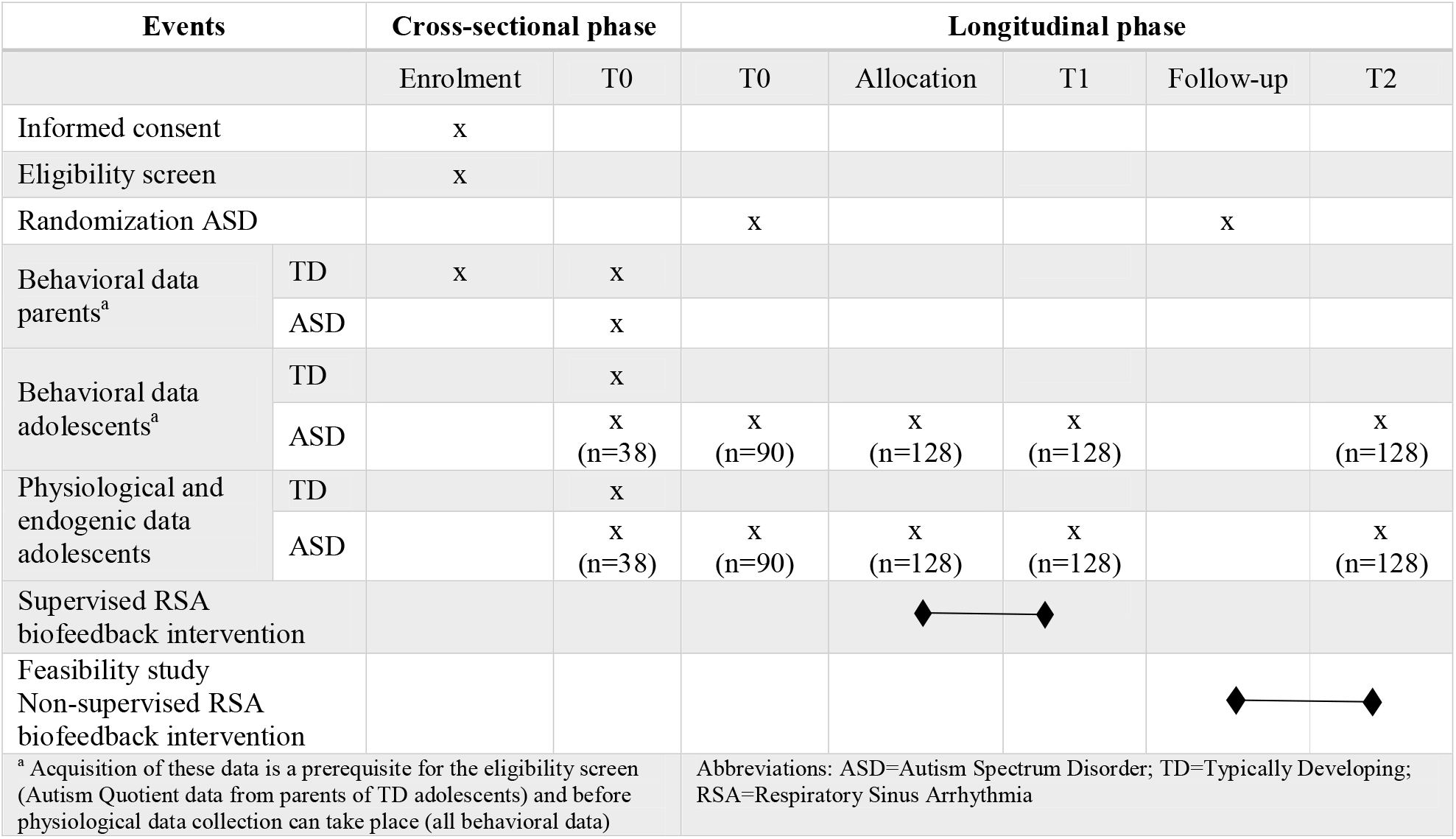
Procedure of enrolment, interventions and assessment following SPIRIT guidelines (Chan et al., 2013). T0 baseline assessment, T1 assessment after five weeks of supervised intervention, T2 after five weeks of follow-up.

The first objective relates to the cross-sectional phase in which adolescents with ASD (n = 38; 19 boys, 13-18 years) and typically developing (TD) peers will be recruited. A standardized stress-provoking assessment will be used to provide insight into differences in autonomic functioning between adolescents with ASD and their TD peers. The primary outcome measure is cardiac vagal modulation, which is hypothesized to be lower in adolescents with ASD. In addition, other physiological and behavioral parameters as well as cortisol measurements will be included as secondary outcome measures. These group comparisons will be highly informative to delineate indices of ANS functioning that are specifically implicated in ASD, and to evaluate whether aberrant physiology in ASD is associated with clinical and behavioral indices (e.g. predictive of increased social difficulties, repetitive behaviors, anxiety…) as reported previously.

The second objective relates to the longitudinal phase in which only adolescents with ASD (n=128) will be included. In this phase, the efficacy of a 5-week supervised RSA biofeedback intervention will be assessed by means of a randomized, single-blinded and sham-controlled design. The intervention is hypothesized to lead to increments of cardiac vagal modulation as opposed to the control intervention. To determine whether the latter hypothesized increment can be maintained, a 5-week follow-up phase will be included. Further, it is hypothesized that an increase in cardiac vagal modulation relates to changes in behavior (e.g. improvement in social behavior) and/or physiological changes but that these might occur at a later stage, thus after the follow-up phase. Therefore, other physiological and behavioral parameters as well as cortisol measurements will be assessed as well.

## Methods

The study is reported conform the SPIRIT (Standard Protocol Items: Recommendations for Interventional Trails) statement (Chan et al., 2013) (see Supplementary Information 1).

### General procedure

The study design is visualized in Table 1 and Figure 1. During the cross-sectional phase, physiological, cortisol and behavioral data will be gathered in adolescents with and without ASD. This phase will also be used as a baseline measurement (T0) for the adolescents with ASD (n = 38) who will subsequently be included in the longitudinal phase. Ninety additional adolescents with ASD will execute the same baseline measurement but their data will only be used for the longitudinal phase. After 5 weeks supervised RSA based biofeedback intervention (or sham-control), all adolescents with ASD (n = 128) will be re-assessed (T1). A follow-up session of 5 weeks will be included (T2) to assess the possibility of retention effects and to determine the presence of behavioral changes. During this follow-up phase, a feasibility study regarding a non-supervised RSA based biofeedback intervention will be conducted by half of the adolescents with ASD (n = 64). All assessments and supervised sessions will be performed at a site from the KU Leuven. In addition, the specific government regulations with regard to COVID-19 will be followed rigorously.

### Ethics approval and informed consent

Ethical approval for this study was granted by both the Ethics Committee UPC KU Leuven on July 2, 2020 (ref. EC2020-541) and the *Ethics Committee Research UZ/KU Leuven on October 20, 2020 (ref: S*64219). Parents of the participants should provide informed consent in addition to the informed assent provided by the participants themselves. Participants may withdraw consent and participation at any time. The participant’s request to withdraw from the study will always be respected and reasons to withdraw are not obliged to be mentioned. The sponsor of this study is KU Leuven.

### Monitoring of side-effects

An independent researcher, who is not involved in data analysis or data collection, will review potential reports of side effects for all participants during and after completion of the study and will be available to discuss side effects with the participants if needed. Standard operating procedures will be followed for reporting serious adverse events to researcher(s), the Ethical Committee and applicable Competent Authorities (CA’s) based on applicable legislation. All participants will be ensured through the no-fault policy of the KU Leuven.

### Participants

A subgroup of adolescents with a formal diagnosis of ASD (n = 38, 19 boys, 13-18 years) based on DSM-IV/5 criteria (American Psychiatric Association, 2000, 2013) will participate in the cross-sectional phase while the total sample (n = 128; 64 boys, 13-18 years) will be included in the longitudinal phase of this study. An age and gender matched group of TD adolescents (n = 38) will be included in the cross-sectional phase only. Exclusion criteria are the presence of an intellectual disability as described in the DSM-IV/DSM-5; the presence of an uncorrected hearing- or vision impairment; the presence of congenital heart diseases, diagnosed cardiovascular abnormalities or somatic diseases with a known impact on heart function; pregnancy; insufficient knowledge of Dutch language and participation in other clinical trials. Use of medication and the presence of comorbid or co-occurring disorders will not lead to exclusion of the study but will be registered elaborately. For TD adolescents specifically, a score equal to or more than 32 on the Autism Quotient – Adolescent Version (Baron-Cohen et al., 2006) and the presence of a neurodevelopmental disorder or psychiatric disorder as described in the DSM-IV/DSM-5 will lead to exclusion from the study. In addition, TD adolescents with a sibling diagnosed with a neurodevelopmental disorder as described in the DSM-IV/DSM-5 will be excluded. These criteria are described in detail in the informed consent and assent forms. If TD adolescents are excluded from the study due to any of the previously described exclusion criteria, they and their parents are encouraged to contact the clinicians associated with this study to discuss their concerns, if any.

Recruitment of adolescents with ASD will take place in the region of Leuven (Belgium) through the Leuven Autism Expertise Centre (a supraregional outpatient clinic supervised by Prof. Steyaert, supporting PI of this study) and the Leuven Autism Research Group (LAuRes, all authors are members). In addition, several contacts have been established with special education schools, autism advocacy organizations and support groups (f.i. VVA, Participate, ‘Autisme Centraal’) as well as with Dr. Klockaerts from the ZNA University Child and Youth Psychiatry in Antwerp and independent clinical practices in and around the region of Leuven. TD adolescents will be recruited in the same region by contacting schools.

### Sample size

Sample size was calculated for the cross-sectional phase using the G*Power tool (Faul F, 2009) with a medium effect size (*g* = 0.59) as reported in the meta-analysis of Cheng et al. (2020), where baseline RSA values were compared between individuals with and without ASD. An a priori power analysis (α=0.05; 1-β=0.80) in a two-independent means design resulted in a total sample size of 74 adolescents, including 37 adolescents with ASD and 37 TD adolescents. In order to achieve an equal gender distribution, the sample size is further increased to 38 adolescents with ASD and 38 TD adolescents.

For the longitudinal phase, there was no evidence for an effect size from a representative study. Therefore, we consider a medium effect size (*d*=0.50) to be representative of a clinical meaningful change. Comparison for the intervention phase was based on the supervised intervention group versus the sham-control group, resulting in an a priori power analysis (α=0.05; 1-β=0.80) for a two-independent means design. The total sample size resulted in a total of 128 adolescents with ASD, considering an hypothetical 20% drop-out rate (Furlan et al., 2009) and an equal gender distribution across both groups (n = 64/group). Since the intervention during the follow-up phase (T1-T2) is a feasibility study, a sample containing 50% of the total sample was deemed to be sufficient in order to obtain reliable information regarding feasibility and acceptability concerning the non-supervised intervention.

### Assessment

As presented in Table 1, informed consent/assent and screening for eligibility will be performed first. In addition, participating adolescents and their parents will be asked to complete the questionnaires prior to scheduling the first study visit.

At each assessment session (T0, T1, T2), a standardized stress-provoking protocol will be executed in which physiological, cortisol and behavioral data will be gathered (see section ‘Outcome Measures’). The collection of behavioral data will differ for the three assessment points as visualized in Figure 1. During the stress-provoking assessment, the participants will be asked to sit with their knees in a 90° angle, with their hands on their thighs, palms facing upwards and eyes closed (during baseline measurement only). To prevent temperature transference of the thighs towards the temperature sensor of the hands, a towel will be placed on the thighs (Laborde et al., 2017). Each of the assessments will be executed in the afternoon, as recommended by Hollocks et al. (2014), to rule out influence of the cortisol awakening response as this can cause misleading results. In addition, participants will be asked to refrain from eating, intense physical activity, smoking and drinking caffeine or alcoholic substances for at least one hour before testing (Smeekens et al., 2015). Following the recommendations of Schmalenberger et al. (2019), female participants will be asked whether they use contraceptives and when their last menstrual period was as these factors influence cardiac vagal activity.

The ‘Social Stress Recall Task’ (SSRT; (Bishop-Fitzpatrick et al., 2017; Richman et al., 2007)) and the ‘Stroop Word-Color Interference task’ (Stroop, 1935) will be used as stress-provoking tasks during the assessments. Both of these measures are validated to provoke stress reactivity as assessed in individuals with ASD (Bishop-Fitzpatrick et al., 2017; Kushki et al., 2013; Richman et al., 2007).

### The ‘Stroop Word-Color Interference task’

A protocol previously used in children with ASD will be used as a guidance (Kushki et al., 2013). Participants will perform a computerized version with five one-minute trials. During the first and the fifth trial, the presentation of words corresponding to color names are printed in the same color as the word itself. The remaining trials consist of incongruent stimuli, meaning that the color name is printed in another color than the word itself. The participants only need to name the color of the print. In addition, the time of the word presentation varies per trial between 1 and 2 seconds. Prior to starting the task, participants are provided with ten practice stimuli to ensure understanding of the task.

### The ‘Social Stress Recall Task’

This task contains a 5-minute stressor phase during which the participant is asked to describe the three most challenging aspects of his/her life on a day-to-day basis. The researcher can use prompts in order to guide the participant such as: “Can you tell me more about how that experience made you feel?” or “Can you remember what you did during that situation?” The timing of prompts will be recorded (Bishop-Fitzpatrick et al., 2017).

### Intervention

The intervention is based on the biofeedback protocol provided by Lehrer (2013) and aims at increasing cardiac vagal modulation in adolescents with ASD as has been demonstrated in previous research (Lehrer et al., 2020).

In order to follow autism-specific treatment guidelines, all training sessions will be summarized and depicted in schedules (Fuentes et al., 2020). This way, the different phases are predictable, and the participants are able to follow each step by themselves. Participants will be asked to provide weekly updates on medication status by visit (during the supervised sessions) or by a reminder mail send by the researcher (during the follow-up phase).

To enhance treatment compliance and gain insight into the feasibility, the participants will be asked – via a secure web-based application (m-Path, KU Leuven) – to rate their level of comfort during the training on a scale using five smiley faces ranging between frustrated and satisfied after each training-session at home.

### Supervised RSA biofeedback

This intervention part lasts for 5 weeks and contains the fixed scheme of one guided session per week with an experienced researcher (AT) and home-practice on days without a guided session. Participants will be asked to breath at their resonance frequency, which refers to the breathing frequency at which heart rate and the breathing pattern are in phase, thus maximizing RSA (Lehrer et al., 2020). During the first session of this intervention, the resonance frequency of the participants will be determined using Biotrace+ Software (Mind Media B.V., www.mindmedia.com), based on the recommendations of Shaffer and Meehan (2020). Subsequently, the participants will be instructed to breath at this frequency through stepwise guidance from the researcher using appropriate breathing techniques (with pursed lips and abdominal breathing) and the provision of a breathing pacer. During home practice, the participants will be provided with a breathing pacer application, individually set to practice at the specified resonance breathing frequency (Awesome Labs LLC, 2020; Trex, 2015). From the third supervised session onwards, participants will be instructed to breath in phase with their heart rate. The latter will be visualized on a computer screen and will replace the breathing pacer installed at their resonance frequency as described for the previous sessions. However, they will continue breathing at resonance frequency during home-practice. The sham-control group will follow the same steps as outlined above but they will practice at a normal breathing pace as determined during the baseline measurement at T0, thus faster than their resonance frequency. In addition, they will not receive any information regarding their own heart rate, as a default mode will be shown during the guided sessions. Each guided session will last approximately 30 minutes with several breaks in between whereas home practice sessions will last 20 minutes, which will be divided into four blocks of five minutes. The participants will be asked – via a secure web-based application (m-Path, KU Leuven) – to rate the level of comfort during home-practice on a scale using five smiley faces ranging between frustrated and satisfied after each session at home.

### Non-supervised RSA biofeedback: feasibility

The non-supervised intervention will be home-based with one guided session at the start during which the individual resonance frequency of the participants will be determined and the procedure will be explained. Next, participants will breath at this resonance frequency on a daily basis in four blocks of five minutes using a biofeedback smartphone application (Elite HRV, Asheville, North Carolina, USA). In addition, a compatible heart rate sensor (Polar^®^ H10, Polar, Finland) will monitor the participant’s heart rate. Information regarding this monitoring will be send to the researcher (AT). A secure web-based application will ask the participants to rate their level of comfort at a daily basis, as described previously in the supervised intervention.

### Randomization

A computer-generated list will be used including block randomization procedures with an allocation ratio of 1:1 for the supervised intervention group versus the sham-control intervention (T0-T1). In addition, a stratification procedure based on gender and severity of ASD (using RBS-R and SRS-2) will be applied in order to obtain an equal division of these characteristics across the intervention groups. Randomization procedures will be performed by AT. Blinding of participants concerning group allocation is only feasible during the supervised intervention, not during the feasibility study with regard to the non-supervised intervention (T1-T2). After completion of the study, participants will be unblinded for group allocation upon request as described in the informed consent/assent.

### Outcome measures

#### Primary outcome measure

##### Cardiac vagal modulation

will be based on RSA, which can be calculated using a time domain measure (Root Mean Square of Successive Differences between normal heartbeats, RMSSD) and a frequency domain measure (Logarithm of High-Frequency Heart Rate Variability, LnHF-HRV) (Laborde et al., 2017). For these parameters, 3-lead electrocardiographic recordings of heart rate (chest placement) will be performed using a NeXus-10 MKII biofeedback device and Biotrace+ Software (Mind Media B.V., www.mindmedia.com) (see subsequent section ‘Physiological parameters’). The recommendations of the Task Force of the European Society of Cardiology and the North American Society of Pacing and Electrophysiology (Malik, 1996) and Laborde et al. (2017) regarding the calculation of this outcome measure will be followed.

### Secondary outcome measures Physiological parameters

#### Heart rate, breathing frequency, skin conductance and fingertip temperature

The NeXus-10 MKII biofeedback device and Biotrace+ Software (Mind Media B.V., www.mindmedia.com) will be used to register the physiological parameters. Continuous measurement during the stress-provoking assessment will be performed with multiple electrodes placed on the body including:

- A 3-lead ECG on the chest with disposable pre-gelled electrodes (Kendall™ ECG Electrodes Arbo™ H124SG, Covidien, Ireland) without specific skin preparations for the recording of heart rate at a sampling rate of 256 SPS.
- An elastic band with stretch-sensitive sensors around the waist for the recording of the breathing frequency at a sampling rate of 32 SPS.
- Two electrodes for the measurement of skin conductance with a sampling rate of 32 SPS strapped around the palmar side of the distal phalanx of the index and ring finger on the non-dominant hand.
- One temperature sensor with a sampling rate of 32 SPS placed around the palmar side of distal phalanx of the middle finger on the non-dominant hand for the recording of fingertip temperature.

Placement of the electrodes is visualized in Figure 2.

**Fig. 2.**
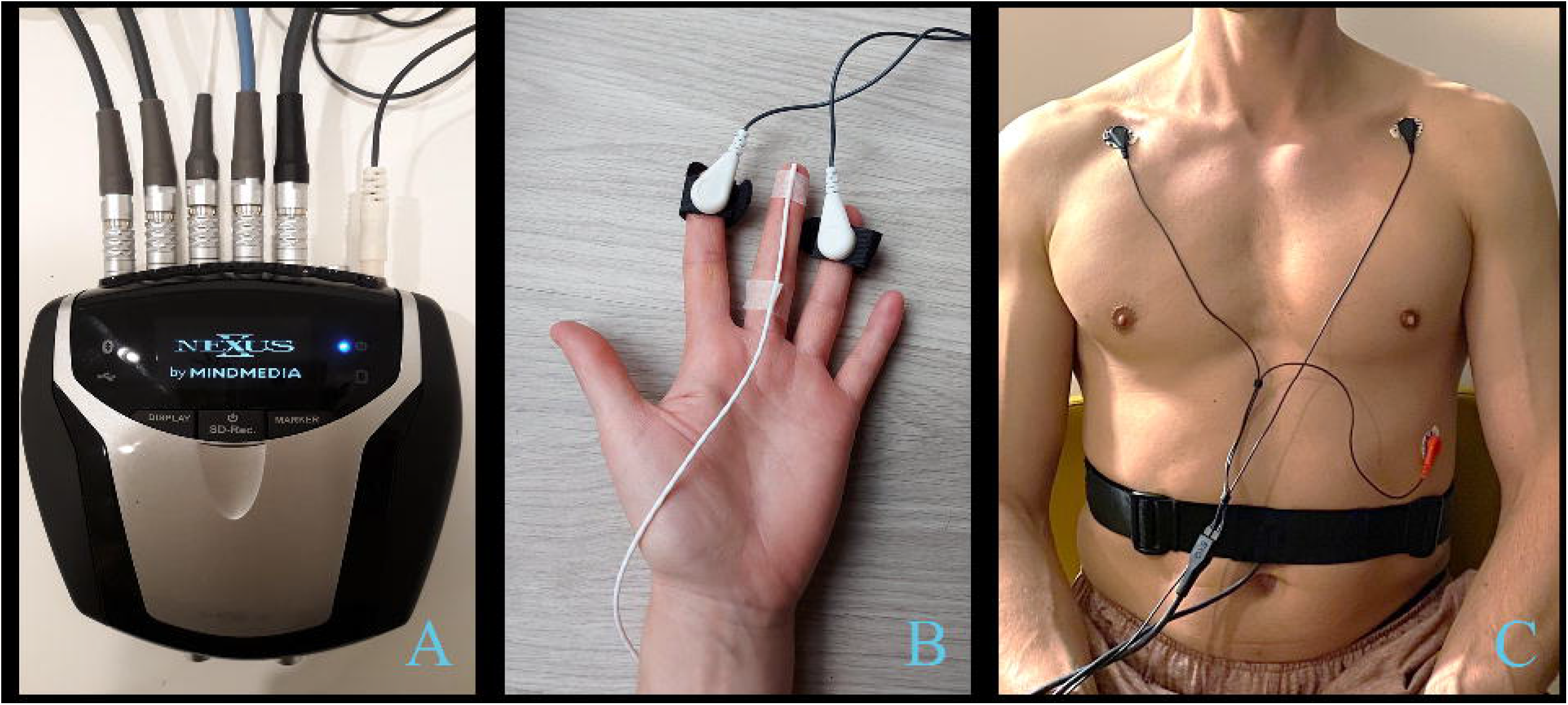
Overview Placement of Physiological Sensors Figures are submitted as separate PNG-files and were created with Microsoft PowerPoint 2016.

#### Cortisol measurement

Salivary samples will be collected to determine the level of cortisol at three time points during each assessment, including a baseline swab and 20 minutes after each of the stress-inducing tasks. Therefore, Salivette^®^ Cortisol cotton swabs (Sarstedt Inc., Rommelsdorf, Germany) will be used. These swabs are validated for use with salivary cortisol. After collection, the swabs will be stored under appropriate conditions (−20°C) at the biobank of the KU Leuven and analyzed in a collaborative lab from the KU Leuven.

#### Behavioral data

##### Autism Quotient – Adolescent Version

The AQ (Baron-Cohen et al., 2006) identifies the extent of autistic traits shown by a person of normal intelligence based on parent-report. A higher total score reflects a higher presentation of autism related symptoms. A cut-off of 32 or higher is used for the exclusion of TD adolescents.

##### Social Responsiveness Scale – Second edition

The SRS-2 (Constantino & Gruber, 2012) is a quantitative assessment based on parent-report. This questionnaire identifies social impairment associated with ASD and quantifies its severity by using five subscales (Social Awareness, Social Cognition, Social Communication, Social Motivation, and Restricted Interests and Repetitive Behavior). Higher scores reflect a higher level of impairment.

##### Repetitive Behavior Scale – Revised

The RBS-R (Bodfish et al., 2000) is used to score the number and intensity of repetitive and restrictive behaviors based on parent-report. A higher total score indicates a higher prevalence of repetitive and restrictive behaviors. In addition, these behaviors are clustered in six categories: Stereotyped Behavior; Self-injurious Behavior; Compulsive Behavior; Ritualistic Behavior; Sameness Behavior. Parents are also asked to rate the impact of these behaviors on daily functioning on a scale from 0 (meaning no interference with daily life) to 100 (extremely problematic related to functioning in daily life).

##### Strengths and Difficulties Questionnaire

The SDQ (Goodman, 1997; Goodman et al., 1998; van Widenfelt et al., 2003) is a brief behavioral screen resulting in five categories of behavioral problems: emotional problems, conduct problems, hyperactivity, peer problems and prosocial behavior. Higher total scores – excluding the prosocial category – indicate more behavioral problems. Both the self-report and parent-report versions will be used.

##### Perceived Stress Scale – Adolescent Version

The PSS (Van der Ploeg, 2013) measures the degree to which situations in the adolescent’s life are appraised as stressful. This measure is based on adolescent self-report in which items are adapted according to the specific problems that adolescents can encounter in their lives, such as school-related stress. A higher total score is related to higher perceived stress levels.

##### Depression Anxiety and Stress Scale – 21-item version

The DASS-21 (Beurs, 2001) measures the presence of negative emotions based on adolescent self-report. Separate scale scores are calculated for symptoms related to depression, anxiety and stress with higher scores reflecting a higher prevalence of the related symptoms.

##### Physical Activity Vital Sign

The PAVS (Greenwood et al., 2010) determines the level of physical activity during the last week based on adolescent self-report. This measure consists of two short questions related to (1) the number of days the adolescent was physically active and (2) the number of minutes (on average) this physical activity was endured.

##### Sensory hypersensitivity based on a Visual Analogue Scale

Measures the self-reported influence of sensory hyper-responsivity on a Visual Analogue Scale going from 0 (meaning no perceived sensory hyper-responsivity) to 100 (meaning presence of severe sensory hyper-responsivity and negative impact on functioning).

##### Perceived stress based on a Visual Analogue Scale

To rate the self-reported perceived stress level on a Visual Analogue Scale per stressor during the stress inducing test. The scale score can vary between 0 (meaning no stress) and 100 (meaning highest level of perceived stress possible).

#### Data handling and statistical analysis

Behavioral data will be gathered via a Qualtrix and m-Path account protected by the KU Leuven. Physiological data regarding heart rate will be exported from BioTrace+ as raw data of all sessions into IBI text files for each participant. These will be exported to Kubios HRV Premium (version 3.4.3, University of Eastern Finland, Kuopio, Finland), which will be used to clean the data using the automatic artefact correction algorithm included in the software. Subsequently, analysis of the data regarding heart rate variability will be performed to determine cardiac vagal modulation expressed as RMSSD and LnHF-HRV, as described in the ‘Primary outcome measure’ section. Researcher-developed scripts in MATLAB R2020b (MathWorks, Natick, Massachusetts, USA) will perform preprocessing and analysis of skin conductance, fingertip temperature and respiration. One member of the research team (AT) will gather all data to conduct statistical analyses using MATLAB R2020b statistical packages to test the main hypotheses. For all analyses, level of significance is set at 0.05 with appropriate corrections for related post-hoc analysis. In addition, the strategy of intention-to-treat will be applied. If normal distribution of physiological data is not present, log-transformation will be conducted as recommended by Laborde et al. (2017). For participants with missing data on any measure, multiple imputation or an ignorability analysis based on the likelihood will be used. If available, reasons for dropout will be listed in order to gain more insight into the feasibility of both interventions.

#### Cross-sectional data comparison

Data from the stress-provoking assessment at T0 will be used to compare the adolescents with ASD (n = 38) with their TD peers (n = 38) in a repeated measures ANOVA with group (ASD vs TD) as between-subject factor. Pearson correlation analysis will be performed to determine correlations between the primary outcome measure and secondary outcome measures.

#### Efficacy of the supervised RSA biofeedback intervention

The efficacy of the supervised intervention will be determined by intention-to-treat analysis using physiological and cortisol data in a mixed model analysis with ‘group’ (intervention vs sham) as a between-group factor and ‘time’ (T0, T1 and T2) as a within-group factor.

#### Change in behavioral outcome measures after supervised RSA biofeedback

In order to determine the influence of the intervention on the behavioral outcome measures, pre-to-post difference scores (T0-T2) will be calculated for the primary outcome measure and the behavioral outcome measures. These differences will be used in a correlation analysis to determine whether positive effects on the primary outcome measure correlate with similar effects on the behavioral parameters.

#### Impact of behavioral outcome measures on treatment efficacy

Mediation analysis will be performed to determine the influence of baseline behavioral characteristics of the participants on the efficacy of the biofeedback interventions. Behavioral parameters from T0 will be split into categories (such as low perceived stress versus high perceived stress) and will be used in split-group-analysis to evaluate pre-to-post differences in the primary outcome measure.

## Discussion

The results of the cross-sectional phase will provide more insight into the differences of autonomic functioning between adolescents with ASD and their TD peers. In addition, the association between autonomic functioning and behavioral measures will be clarified. The longitudinal phase will determine the efficacy of a supervised RSA biofeedback intervention in adolescents with ASD. This intervention may provide multiple benefits for this population specifically due to its possibility to be practiced at home, as will be examined during the feasibility study of the non-supervised RSA based biofeedback intervention in this study. The latter can facilitate the integration and transfer into their daily routine. Given the high rate of co-occuring psychological symptoms and a hypothesized association between lower cardiac vagal modulation and these symptoms in individuals with ASD, an upregulation of cardiac vagal modulation may be valuable for this particular population. This can be accomplished by RSA based biofeedback intervention, as has been previously demonstrated in other populations including physical, behavioral, and cognitive conditions (Chudleigh et al., 2019; Darling et al., 2019; Gevirtz, 2013; Goodman et al., 2018; Kennedy & Parker, 2019; Knox et al., 2011; Lehrer et al., 2020; Lehrer & Gevirtz, 2014; Thabrew et al., 2018).

### Study status

This study was registered at ClinicalTrials.gov on November 13^th^, 2020 (NCT04628715). At the time of initial manuscript submission, recruitment had started (December 2020) but not yet completed.

## Data Availability

This is a protocol paper thus no data is available.

## Acknowledgements

The research reported was supported by funding from the Marguerite Marie Delacroix Foundation (awarded to AN).

## Abbreviations

ASD: Autism Spectrum Disorder
AQ: Autism Quotient
DASS: Depression Anxiety and Stress Scales
DSM-IV/V: Diagnostics and Statistical Manual of mental disorders
ECG: Electrocardiogram
FU: Follow-up
HF-HRV: High Frequency Heart Rate Variability
HRV: Heart Rate Variability
PAVS: Physical Activity Vital Sign
PSS: Perceived Stress Scale
RBS-R: Repetitive Behavior Scale – Revised
RMSSD: Root Mean Square of Successive Differences
RSA: Respiratory Sinus Arrhythmia
SDQ: Strengths and Difficulties Questionnaire
SRS: Social Responsiveness Scale
SSRT: Social Stress Recall Task
TD: Typically Developing
VAS: Visual Analogue Scale.

